# Impact of essential workers in the context of social distancing for epidemic control

**DOI:** 10.1101/2020.05.05.20092262

**Authors:** William R. Milligan, Zachary L. Fuller, Ipsita Agarwal, Michael B. Eisen, Molly Przeworski, Guy Sella

## Abstract

Many governments have responded to the ongoing COVID-19 pandemic by imposing social policies that restrict interactions outside of the home, resulting in a large fraction of the workforce either working from home or not working. However, to maintain essential services, a substantial number of workers are not subject to these limitations, and maintain many of their pre-intervention interactions. To explore how interactions among such “essential” workers, and between essential workers and the rest of the population, impact disease risk and the effectiveness of pandemic control, we evaluated several models of essential worker interactions within a standard epidemiology framework. The models were designed to correspond to key characteristics of, respectively, cashiers, factory employees, and healthcare workers. We find in all three models that essential workers are at substantially elevated risk of infection compared to the rest of the population, and that increasing the numbers of essential workers necessitates the imposition of more stringent interaction controls on the rest of the population in order to manage the pandemic. However, different archetypes of essential workers differ in both their individual probability of infection and impact on the broader pandemic, highlighting the need to understand and target for intervention the specific risks faced by different groups of essential workers.

## Introduction

In response to the ongoing COVID-19 pandemic, governments across the world have imposed measures to reduce the transmission of the virus. These measures often involve some form of “shelter in place” (SIP) in which the majority of the population remains in their homes except for essential activities like grocery shopping. The motivation is to either locally eradicate the infection, or to reduce its spread enough to decrease peak demand on healthcare and gain time to develop testing capacity, therapies and a vaccine. SIP orders have been guided by extensive modeling of the COVID-19 pandemic to predict its future and understand its impact on the population under various scenarios, including different stringencies of SIP (e.g.,[1–3]).

By necessity, SIP involves exceptions for “essential workers”, typically including those involved in the delivery of health care, the production and distribution of food, emergency services and defense, public works and utilities, communications and information technology, and logistics and delivery. The fraction of workers designated as essential varies geographically due to different regulations and the makeup of the local economy [4]. Within the United States, industries designated as essential are estimated to employ approximately 40 percent of the workforce [4]. In New York City, essential workers were estimated to comprise a quarter of the workforce [5], or over 1M people, of whom over half are employed in healthcare and 15 percent in grocery, convenience and drug stores. Estimates in California are that one in eight [6] individuals is an essential worker.

Here we extend the widely-employed “SEIR” epidemiological model [1–3] to evaluate the individual infection risk faced by different types of essential workers, and the impact that essential workers have on pandemic control.

## Modeling Essential Workers

We began by implementing a standard SEIR model to describe the dynamics of COVID-19 in a population (Figure 1A). Following recent work [1–3], we included three types of infected individuals, corresponding to those destined to a) show no symptoms, b) to have symptoms but not require hospitalization, and c) to require hospitalization. We also included three types of hospitalized individuals corresponding to a) those who will recover without critical care, b) those who will require critical care but will recover, and c) those who will die after receiving critical care. We make the standard modeling assumption that recovered individuals cannot be reinfected.

**Figure 1:**
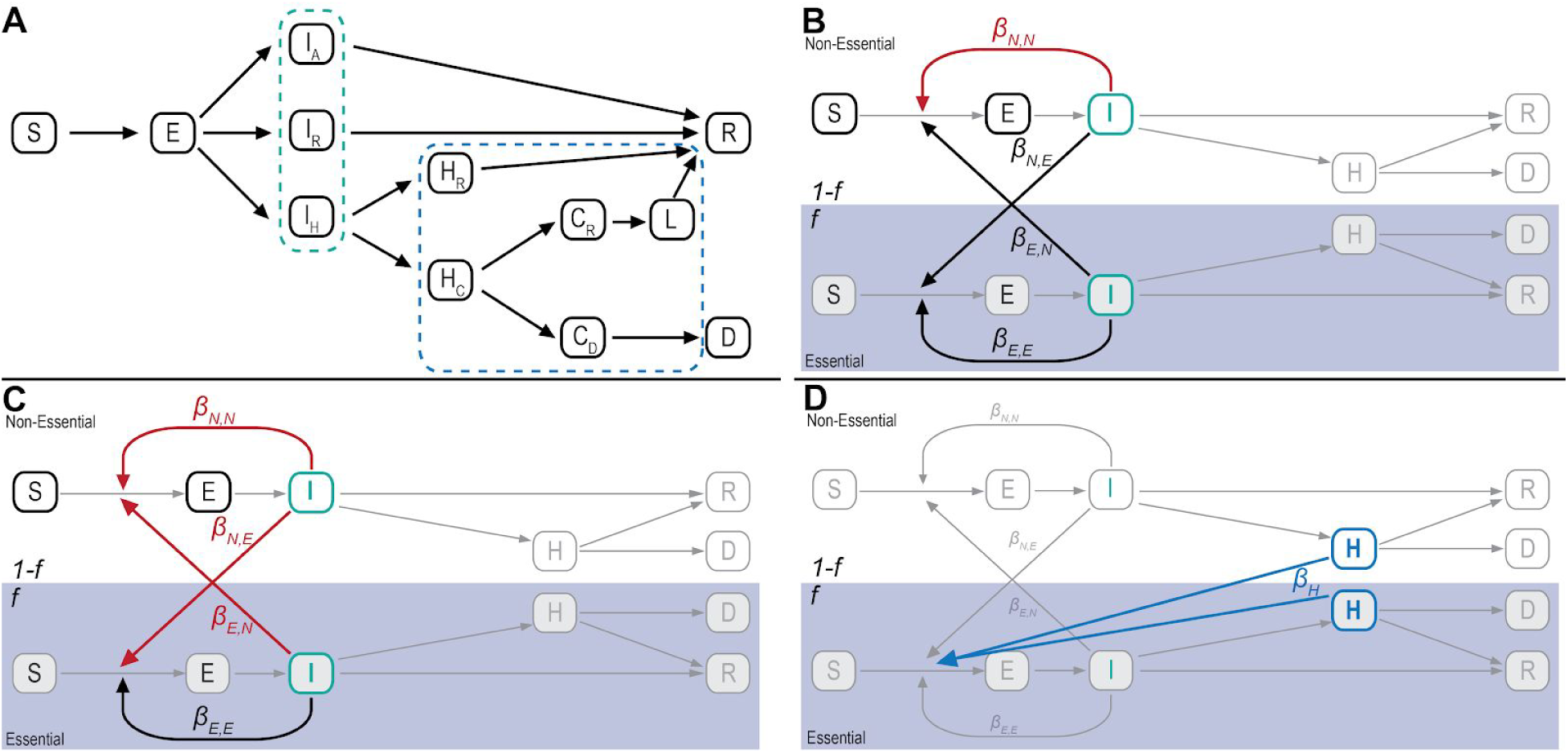
Diagram of the SEIR model with extensions. **A**) An illustration of the basic SEIR model used in all scenarios, including additional compartments within the infected (outlined in green) and hospitalized (outlined in blue) classes. ‘S’ is susceptible, ‘E’ is exposed, ‘I’ is infected, ‘H’ is hospitalized, ‘D’ is dead, and ‘R’ is recovered. Within the infected class, individuals can be asymptomatic ‘I_A_’ and destined to recover; symptomatic ‘I_R_’ but destined to recover; or symptomatic ‘I_H_’ and destined to be hospitalized. Within the hospital, individuals either go to recovery ‘H_R_’ or go to critical care ‘H_c_’. For those in critical care, individuals either die ‘C_D_’ or go on to the recovered class ‘C_R_’, with an additional time spent in the hospital ‘L’. **B**) To model the impact of EWs, who make up a proportion ***f*** of the total population, we cloned the SEIR model. Here, for visualization, the infectious and hospitalized classes are collapsed into a single compartment. The *β* terms represent the transmission routes between infectious individuals within and between essential ‘E’ and non-essential ‘N’ groups. In the model of public-facing EWs, SIP reduces interactions (highlighted in red) only among nEWs. **C**) In the model of non-public-facing EWs, SIP reduces all interactions (highlighted in red) except for those among other EWs. **D**) To model the impact of healthcare workers, an additional infectious route (*β_H_*) is included from within the hospitalized compartments to susceptible individuals in the essential group.

We parameterized the differential equations that describe the rate at which individuals transition between model states based on the epidemiological literature for COVID-19 (see Table 1), and chose the lockdown date and *R*_0_ (the number of new infections expected to arise from an infected individual in a fully susceptible population) based roughly on New York City data (see Modeling Details). Although it was not our goal to model the outbreak in any particular region, we verified that the parameters we used match observed pandemic dynamics in three US cities (see the computational notebook).

**Table 1:** Parameter estimates for the model

| Parameter | Description | Literature Estimates | Values Used |
| --- | --- | --- | --- |
| $t_E$ | Latency period | 4.6 days [1,2]<br>5 days (1-14) [24] | 4.6 days |
| $t_{IA}$ | Infectious period (asymptomatic class) | 5 days[1,2]<br>8 days [24] | 5 days |
| $t_{IR}$ | Time of between infection and recovery (mild symptoms) [infectious period for mild cases] | 5 days [1,2]<br>8 days [24]<br>Median 2 weeks [25]<br>1-2 weeks [26] | 5 days |
| $t_{IH}$ | Time between infection and hospitalization | 5 days [1,2]<br>1 week [25]<br>5-12 days [27]<br>7 +/- 4 days [28] | 5 days |
| $t_{HR}$ | Time in hospital (with no critical care) | 8 days [1,2]<br>11 days [24] | 8 days |
| **Total duration of infectious period through critical care path (to recovered or dead)** |  | 21 days [1,2]<br>3-6 weeks [25]<br>16-23 days [28] | 21 days |
| $t_{HC}$ | Time in hospital before critical care | 6 days[1,2] | 6 days |
| $t_{CR}$ | Time in critical care before recovery | 10 days[1,2] | 7 days |
| $t_L$ | Time in critical care before recovery | - | 3 days |
| $t_{CD}$ | Time in critical care before death | 10 days [1,2] | 10 days |
| $p_{EIA}$ | Proportion of infections that are asymptomatic | 33% [1,2] 17.9% [29]<br>13.5% [30] 25% [31] | 33% |
| $p_{EIH}$ | Proportion of infections requiring hospitalization | 4.4% [1,2] | 4.4% |
| $p_{EIR}$ | Proportion of symptomatic not requiring hospitalization | - | 62.6% |
| $p_{IHR}$ | Proportion of hospitalizations requiring critical care | - | 30% |
| $p_{IHC}$ | Proportion of hospitalizations that require critical care | 30%[1,2]<br>26%-32% [27] | 30% |
| $p_{HCR}$ | Proportion of recoveries from critical care | - | 50% |
| $p_{HCD}$ | Proportion of deaths in critical care | 50% [1,2]<br>39% to 72% [27] | 50% |

We next created two parallel instances of the base SEIR model, one for essential workers and one for everyone else, and connected them via terms that describe the probability that an infected individual in one subpopulation infects a susceptible individual in the other (Figure 1B-D). In what follows we refer to these two subpopulations as essential workers (EWs) and non-essential-workers (nEWs), the latter category encompassing all other workers and people not in the labor force. The models for EWs and nEWs have identical structure and parameters, except for the within and between subpopulation transmission rates.

For our analysis, the critical characteristic of EWs that distinguishes them from nEWs is that they maintain a substantial fraction of their work-associated interactions after the institution of SIP. However, there is great diversity among EWs in their interaction profiles. For example, factory, warehouse and agricultural workers retain interactions with the other employees at their place of work, but have their interactions with the remainder of the population reduced by SIP. Others, such as cashiers, transportation workers, and police have frequent interactions with many people who are nEWs. Hospital workers, in turn, interact not only among themselves, but also with the people hospitalized with the virus.

We therefore generated three separate EW-containing SEIR models, one for each of these archetypal EWs. These models differ in how an individual’s interactions are distributed within and between subgroups, in how SIP affects these interactions, and, for healthcare workers, in which individuals are the source of new infections.

The three models have two shared parameters: *f*, the fraction of the population that are EWs, and θ, the remaining proportion of individual to individual disease transmission after social distancing (θ = 1 is no social distancing; θ = θ is complete isolation for everyone). Without EWs (*f* = 0), the pandemic is suppressed when *R*_0_θ (the post-SIP number of new infections expected to arise from an infected individual in a fully susceptible population) is less than one.

Mathematical details and full parameter choices for each model are described in the Modeling Details section, and implementation and the results of simulations with these models are available in the included computational notebook.

### Model 1: Public-facing essential workers (Figure 1B)

We began by considering workers such as cashiers and other shopworkers, transportation workers and public safety personnel, whose work involves extensive interactions with nEWs. The critical feature of our model of such “public-facing” EWs is that only interactions among nEWs are reduced by SIP.

### Model 2: Non public-facing essential workers (Figure 1C)

Unlike public-facing EWs, factory, warehouse, and agricultural workers interact extensively with other EWs, but their work does not involve interactions with nEWs. The critical feature of our model of such “non-public-facing” EWs is that all interactions except those among EWs are reduced by SIP.

### Model 3: Healthcare workers (Figure 1D)

Healthcare workers are exposed to infected individuals in hospitals and other critical care settings. We therefore created a specific “healthcare worker” model with an additional interaction term describing the rate at which individuals hospitalized with COVID infect susceptible healthcare workers (see Modeling details). Neither these hospital-specific infections nor infections among healthcare workers are affected by SIP, while interactions between healthcare workers and nEWs, as well as interactions among nEWs, are.

## Results

### Essential workers have elevated infection risk

We began by examining the personal infection risk of each class of EW. Figure 2 shows the cumulative fraction of EWs and nEWs infected as the pandemic progresses for each model, with *f =* 0.05 (five percent of the population EWs) and θ value corresponding to partially effective (*R*_0_θ = 1.5), effective (*R*_0_θ *=* 0.9) and highly effective (*R*_0_θ *=* 0.5) SIP.

**Figure 2:**
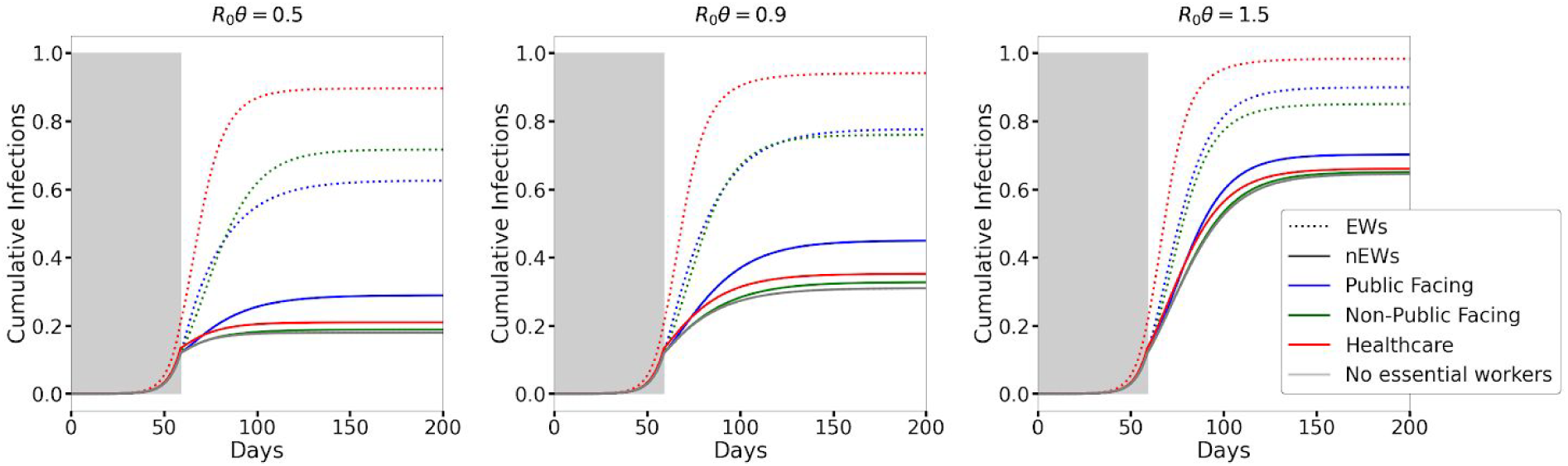
Cumulative infection rates among EWs and nEWs for different scenarios and values of θ. The dotted and solid lines correspond to EWs and the rest of the population, respectively. Note that the ordering of the colors is not the same for EWs and nEWs. *f* is assumed to be 0.05 for all models. Alternative values of *f* yield similar qualitative results, as does an earlier lockdown date relative to the pandemic progression (see the computational notebook).

For all three models, EWs have a substantially higher risk of infection than nEWs. The risk is greatest for healthcare workers, who, under our model parametrization, are nearly all infected quite rapidly, even when the rest of the population is under stringent SIP. But both public-facing and non-public-facing EWs have much higher risk than nEWs: non-public-facing EWs are susceptible to the wave of infection that sweeps through their workplaces even after SIP, while public-facing EWs are exposed to infected people at a much higher rate than those under SIP.

Our model predicts a somewhat higher rate of infection at 100 days than seems plausible, likely because we are not taking into account measures taken by EWs, in particular healthcare workers, to reduce transmission in their workplaces.

### Public-facing essential workers increase infection risk in the remainder of population

Although the individual risk to public-facing EWs is relatively low, they have a substantial impact on nEWs. In conditions when SIP would be expected to be effective at controlling the growth of the pandemic (*R*_0_θ < 1), having five percent of the population working in public-facing EW jobs leads to nearly doubling the number of nEWs who are infected (Figure 2).

We were initially surprised that the high rate of infection of both non-public-facing and healthcare EWs led only to marginal increases in the infection risk for nEWs (Figure 2).

However, the combination of the model assumptions of a relatively small EW population (five percent of the total), that half of EW interactions are with other EWs (see Model Details), and SIP suppression of interactions between EWs and the rest of the population means that, even when there is rampant infection among EW, there is a low leakage of infections to nEWs. And, although some transmission from EWs to nEWs does occur, because of SIP, those infections do not take root in the nEW population.

### Effects on pandemic control of increasing number of essential workers

The differing effects of EWs on disease risk in EWs and nEWs led us to next examine how the different types of EW impact the pandemic. Figure 3 shows the total fraction of the population expected to be infected after one year as a combined function of *f* and θ for all three EW models (Figure 3A), as well as the breakdown for EWs (Figure 3B) and nEWs (Figure 3C).

**Figure 3:**
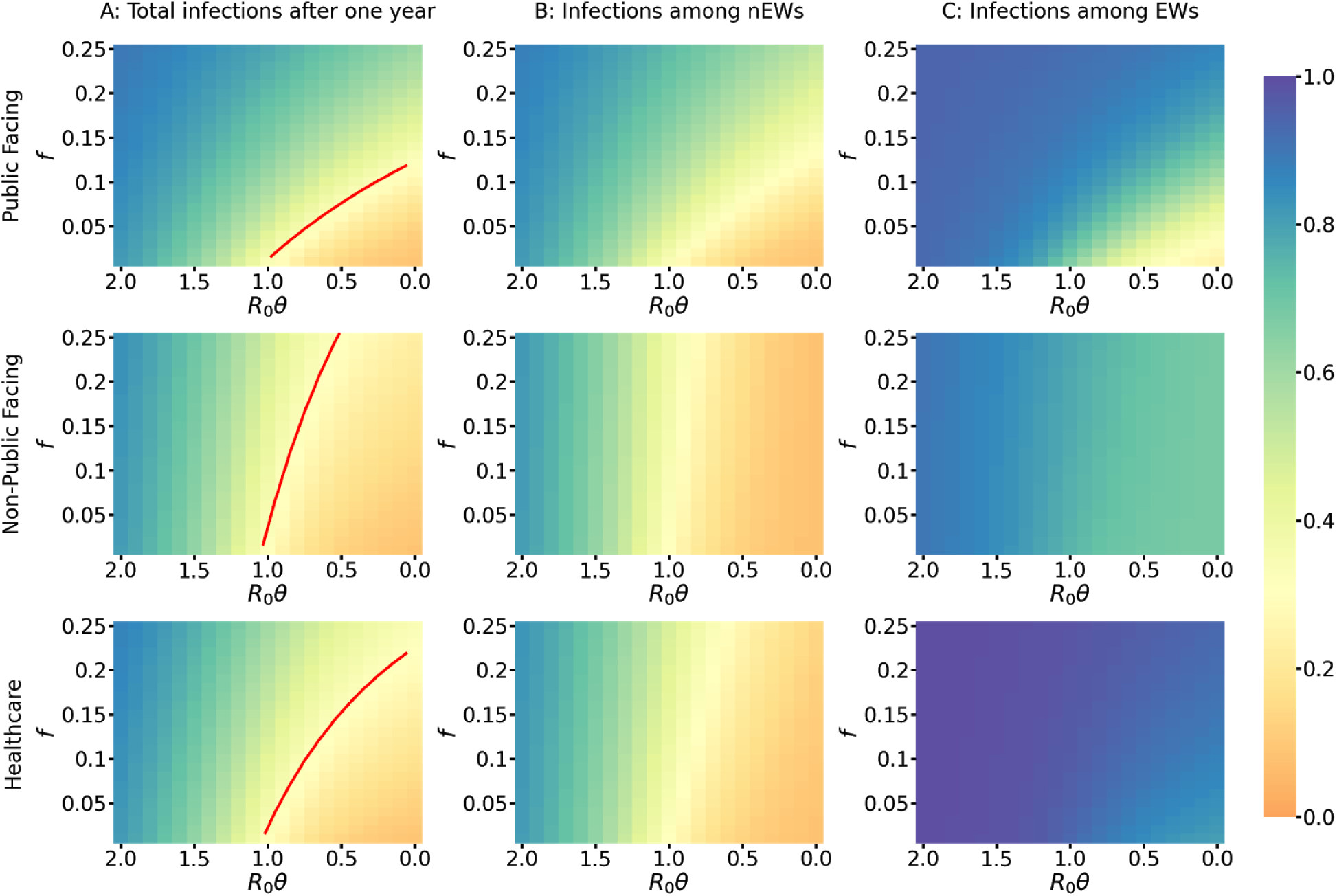
Heatmaps of cumulative infections after a year in the total population (left), in nEWs (middle), and in EW (right). The red contour line corresponds to the number of cumulative infections within a year assuming no EWs. For equivalent figures with an earlier lockdown date see the computational notebook.

The red contour line in each panel of Figure 3A represents *f*, θ values for which the total fraction of the population infected after one year is equal to the fraction infected for *f* = θ and *R*_0_θ = 1. Values below and to the right of this band result in fewer people infected, values above and to the left more.

In all models, increasing the number of EWs requires a compensatory increase in the stringency of SIP. However, there is considerable difference in the stringency required in each case. An approximately two-fold greater decrease in *R*_0_θ is required to compensate for an increase in the number of public-facing EWs compared to the same increase in the number of non-public-facing and healthcare EWs.

### Dynamics of infections to, from and within essential worker populations

The differences in infectious interactions within and between subpopulations after SIP results in a complex dynamics of the number and source of infections of EW and nEW over the course of a year. To investigate their nature, and understand how they manifest in the three different EW models, we examined the prevalence of infection in EWs and nEWs at *f =* 0.05 and *R*_0_θ = 0.5 as a function of time (Figure 4A).

**Figure 4:**
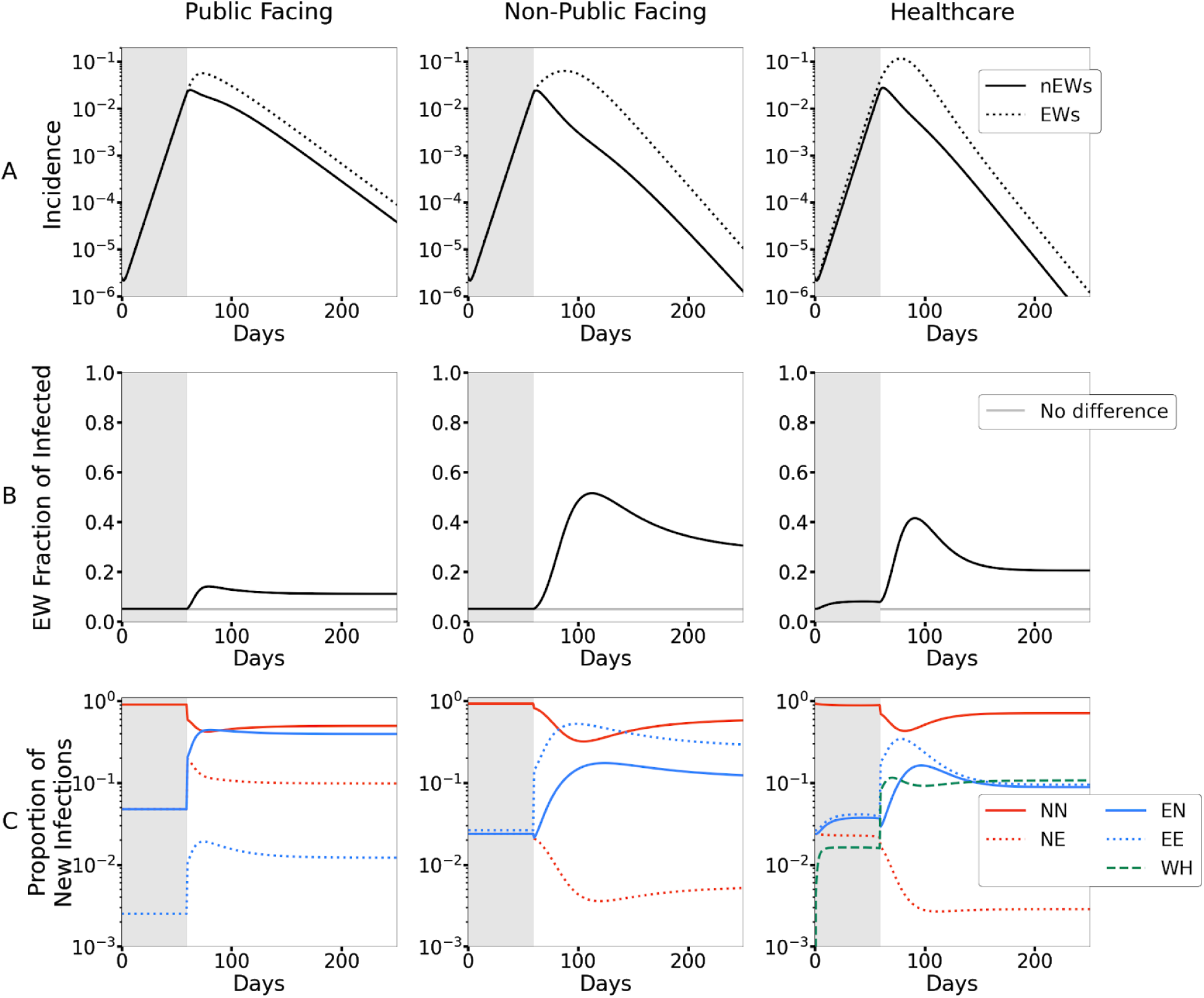
Time resolved dynamics of infections, for *f*=0.05 and *R*_0_θ = 0.5. **A**) Time-resolved proportion of EWs and nEWs that are infected. Before SIP, the epidemic progression is identical in public facing and non-public facing EWs, and in nEWs. However even before SIP, healthcare workers bear a proportionally larger burden of the epidemic due to within-hospital infections. After SIP, the fraction of nEWs infected begins to decline, but, because EWs cannot social distance as effectively as nEWs, the fraction of EWs continues to increase, albeit more slowly. **B)** The fraction of infected individuals that are EWs. Prior to SIP, this fraction is *f* except in the healthcare model where EWs have an additional route of infection. After SIP, EWs bear a proportionally larger burden of the epidemic and this fraction increases. Under the public-facing worker model, the rate of contact between EWs and nEWs isn’t reduced by SIP, mitigating divergence in the rate of infection in the two subpopulations. In the non-public facing and healthcare worker models, SIP largely decouples the epidemic between EWs and nEW. Thus, the increased infection rate among EWs does not cause substantially more infections among nEWs, resulting in EWs comprising a disproportionately large proportion of the infections (i.e., >> *f*). As the epidemic dies out in both subpopulations, the proportion of EWs among those infected stabilizes on a value that is model and parameter-dependent. **C)** Where new infections originate. SIP reorganizes the flow of infections through the populations in a model-dependent manner. Before SIP, the most infections spread among nEWs, as they constitute the vast majority of the population. After SIP, we would naively expect a reduction in infections stemming from interaction terms that are reduced by SIP. Indeed, immediately after SIP, infections associated with reduced interaction terms are reduced (e.g. EN in non-public facing and NN in all models). However, EWs also bear a disproportionately large burden of infections. Thus after a short lag, as infections among EWs increase, so does their contribution to new infections in both other EWs and in nEW; in fact EWs often become the predominant spreaders of new infections. For equivalent figures with a lockdown earlier in the pandemic progression, see the computational notebook.

Unsurprisingly, after SIP is imposed, the incidence of infections in nEWs begins to decline, while the prevalence in EWs continues its previous rise. In all three models, infections in EWs rapidly peak, after which EW prevalence tracks with nEW prevalence, albeit at a higher level. There is, however, a striking difference between the public-facing EW model and the other two, with both a slower decay of incidence, and less of a gap between EWs and nEWs in the public-facing EW model.

This difference is more evident when examining the fraction of all infections that are in EWs as a function of time (Figure 4B). In the public-facing EW model, there is a rapid rise from the initial setting 0.05 to a peak of 0.15, where it levels off. In contrast both the non-public facing and healthcare EW models reach a point where roughly half of all infections occur in EWs before stabilizing slowly to a value of approximately one third.

The changes in the distribution of infections across the EW and nEW populations over time results in complex, shifting patterns of who is infecting who (Figure 4C). In the public-facing EW model, there is a rapid shift post-SIP to there being a roughly equal probability that a new nEW infection came from either another nEW or an EW - a striking result given that in this scenario only five percent of the population are EWs. In contrast, public-facing EWs are roughly ten times more likely to be infected by an nEW than an EW.

The non-public-facing EW model has quite different dynamics. It takes longer for EWs to become a significant source of new infections for nEWs, a product of the time it takes for the infection to spread extensively within the EW population. In contrast to the public-facing model, where EW to nEW and nEW to nEW are effectively tied for the most common type of infection transfer, in the non-public-facing model, the two most common infection types are both within group terms. The dynamics of the healthcare EW model are largely similar to the non-public-facing EW model, except that infections associated with hospital care of infected individuals become a major source of infection for healthcare EWs.

Thus it is a fundamental aspect of all three EW models that one important and potentially observable feature of the pandemic - who is infecting who - is expected to change over time.

## Discussion

It may be intuitively obvious that, when populations are sheltering in place to reduce virus transmission, EWs whose jobs require them to maintain interactions with each other and/or the public have a higher risk of infection, and, if infected, an increased probability of spreading. Yet the precise nature of this effect has received relatively little attention in COVID-19 modeling and public planning. Our goal here was to address a critical issue, how variation in the interaction profiles of common types of EWs affect their disease risk and efforts to control the pandemic, within the context of the epidemiological models that are being widely used to guide policy decisions.

We want to emphasize that, while our modeling has led to several general observations about the potential interaction of EWs with COVID-19, it was not designed to be used for predicting pandemic progress in specific populations with given types and numbers of EWs, different starting states and disease parameters, and distinct types and timings of SIP. This would require, at a minimum, accurate data on interactions among EWs and between EWs and nEWs in the specific context being modeled, incorporation of demographic differences in EW and nEW populations, and treatment of the compartmentalization of both populations into specific workplaces and households [5,7–9].

Moreover, the **R_0_** value we used in the model may be more representative of urban areas, which, because of population density, public transportation and other factors, tend to have a higher rates of contact than do regions with lower population density [10]. Nonetheless, empirical data on rates of infection and fatalities in groups of EWs in urban areas hard-hit by COVID-19 paint a clear picture of increased individual risk [11–14], and the models developed here suggest that, at a minimum, this is likely to be a pervasive challenge in population centers. There is also evidence for elevated EW risk in factories and food-processing facilities in areas with lower population density [15]; however we did not explore how the models behave in such conditions.

We also have not fully explored the full range of scenarios that our models could capture. In particular, we focused on the impact on and of EWs within the context of SIP orders applied in the midst of a rapidly growing local outbreak, as has occurred repeatedly in past few months across the planet. EWs may have a very different disease risk and impact on the epidemic when SIP is applied before a large number of individuals are infected, or after the outbreak is already under control. There is also likely to be a significant effect of the conditions under which EWs under consideration live and work (e.g.,[16]). Our models, for example, do not take into account protective measures taken by EWs, in particular healthcare workers.

What is clear, however, is that the type of essential work in which a person is engaged has a big effect on their individual risk of infection. For example, even with limited exposure to the public, EWs at high interaction workplaces such as manufacturing and food processing facilities, or with high exposure to infected individuals, are at the highest individual risk of infection. But more public-facing workers, such as cashiers, even when they have a much lower individual risk, can have a much greater impact on the pandemic.

Although the specific observations are tentative in their dependence on the assumptions of our models, these results highlight the importance of not treating EWs as an undifferentiated class, and establish the extent of EW interaction with nEWs as a critical feature in studying EWs in the future and designing strategies to mitigate individual and collective risk arising from their essential work.

Our modeling results also emphasize that increasing the number of EWs, as is now being considered worldwide [17-19], will require the remainder of the population to increase the stringency of their shelter in place if pandemic suppression is to be maintained. Thus it is particularly important that policy decisions regarding how and when to expand the EW pool factor in the possible impact of these policies on the willingness and ability of the nEWs to increase or even maintain the stringency of SIP.

Finally, while we treat the interaction profiles, and therefore disease transmission probabilities, of EWs as static, this is by no means the case. Workplace social distancing policies, design changes and the deployment of personal protective equipment in both EWs and nEWs can play a significant role in limiting EW exposure to infected individuals, and, if infected, minimizing their role in disease transmission. Widespread workplace testing for infections and temporary removal of infected individuals from the workplace would also reduce transmission. We hope that modeling efforts like ours can help inform the best way to target these interventions.

## Modeling Details

We were guided by two principles in designing our models for different types of EWs. First, we wanted them to capture essential characteristics of archetypes of EWs: cashiers interact extensively with the public, factory workers have a large fraction of their interactions in the workplace, and healthcare workers are exposed to a unique infection risk from their exposure to a high concentration of infected individuals. Second, we wanted the models to be simple enough that we could connect the modeling results back to these essential characteristics. Hence none of these models should be considered to fully represent a real group of EWs; rather they represent distinct essential characteristics that are often found in real groups of EWs.

All models are described in terms of the number of potentially infectious contacts per day, where these interactions are apportioned between subpopulations according to *β_tj_*, which represents the number of contacts an individual in population *i* has with individuals in population *j*. Representing EWs by *E* and nEWs by *N*, the four corresponding parameters are β*_NN_*, β*_NE_*, β*_EN_*, and β*_EE_*.

We assume that contacts are symmetric, such that with proportion *f* of EWs and 1 − *f* of NEs:

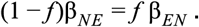

We further assume that *before SIP* every individual has a set total number of contacts β*_T_* such that:

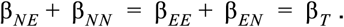

For public-facing EWs, we assumed that before SIP interactions between subpopulations are proportional to the susceptible subpopulation’s size such that:

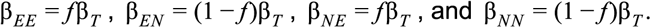

As we are further assuming that public-facing EWs interactions are unaffected by SIP, **θ** is applied only to β*_NN_*, such that after SIP the parameters are:

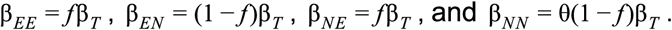

For non-public-facing EWs, prior to the proportional assortment of interactions, we reserve a fraction ρ of their interactions to be with other EWs. Thus, we assume that before SIP:

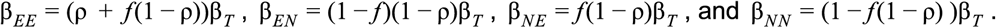

For non-public-facing EWs, we assume that all interactions except the ρβ*_T_* reserved to be among EWs in the workplace are reduced by SIP, and thus post SIP the parameters are:

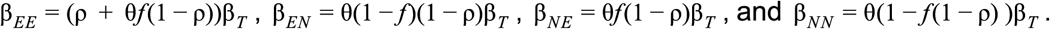

For the examples presented here, we set ρ to 0.5. Other values are explored in the accompanying computational notebook.

The healthcare model builds on the non-public facing model, but in this case all hospitalized individuals have an extra β*_HE_* interactions with healthcare workers per day. We assume β*_HE_* scales with the number of healthcare workers, and parameterize the choice of its value such that a healthcare worker is *κ* times more likely to get infected by a nEW than is a nEW (see the computational notebook for details). In the main text, we set *κ*=1.5. Alternative parameter values and parameterization of β*_HE_* are explored in the computational notebook.

The other parameters used in the SEIR model come largely from [1,2], as detailed in Table 1, with further consideration of the dynamics of COVID-19 in New York City before SIP. In particular, the observed doubling time of three days in the number of deaths in New York City before formal SIP orders [20,21] is best fit with *R*_0_ = 3.2. We used this value to determine the value of β*_T_*. We further placed the start of SIP at day 59, such that our model without EWs predicts approximately the observed number of deaths in New York City on the day formal SIP orders were issued. This is relatively late in the pandemic progression, in that a substantial fraction of the population was already infected (as seems to have been the case in New York City, according to recent serological estimates [22,23]), and accounts for the high prevalences in the models. Similar qualitative results are seen for earlier SIP dates (see the computational notebook).

## Data Availability

A computational notebook is freely available to reproduce all figures and analyses in the text

https://github.com/zfuller5280/Covid-19_Modeling

## Acknowledgments

We thank Laura Hayward, Magnus Nordborg and Michael Zeitz for helpful discussions.

